# Successful Detection of Delta and Omicron Variants of SARS-CoV-2 by Veterinary Diagnostic Laboratory Participants in an Interlaboratory Comparison Exercise

**DOI:** 10.1101/2022.11.08.22282084

**Authors:** Kaiping Deng, Sarah M. Nemser, Kirstin Frost, Laura B. Goodman, Hon S. Ip, Mary Lea Killian, Jodie Ulaszek, Shannon Kiener, Matthew Kmet, Steffen Uhlig, Karina Hettwer, Bertrand Colson, Kapil Nichani, Anja Schlierf, Andriy Tkachenko, Megan R. Miller, Ravinder Reddy, Gregory H. Tyson

**Author notes:** Corresponding author: Gregory Tyson, 8401 Murkirk Road, Laurel, MD 20708 USA.

## Abstract

**Background:** Since the beginning of the COVID-19 pandemic veterinary diagnostic laboratories have tested diagnostic samples for SARS-CoV-2 not only in animals, but in over five million human samples. An evaluation of the performance of those laboratories is needed using blinded test samples to ensure that laboratories report reliable data to the public. This interlaboratory comparison exercise (ILC3) builds on two prior exercises to assess whether veterinary diagnostic laboratories can detect Delta and Omicron variants spiked in canine nasal matrix or viral transport medium.

**Methods:** Inactivated Delta variant at levels of 25 to 1,000 copies per 50 μL of nasal matrix were prepared for participants by the ILC organizer, an independent laboratory, for blinded analysis. Omicron variant at 1,000 copies per 50 μL of transport medium was also included. Feline infectious peritonitis virus (FIPV) RNA was used as a confounder for specificity assessment. A total of 14 test samples were prepared for each participant. Participants used their routine diagnostic procedures for RNA extraction and real-time RT-PCR. Results were analyzed according to International Organization for Standardization (ISO) 16140 - 2:2016.

**Results:** The overall results showed 93% detection for Delta and 97% for Omicron at 1,000 copies per 50 μL (22-200 copies per reaction). The overall specificity was 97% for blank samples and 100% for blank samples with FIPV. No differences in Ct values were significant for samples with the same virus levels between N1 and N2 markers, nor between the two variants.

**Conclusions:** The results indicated that all ILC3 participants were able to detect both Delta and Omicron variants. The canine nasal matrix did not significantly affect SARS-CoV-2 detection.

**Impact Statement:** Ensuring accurate detection methods for SARS-CoV-2 is critical as veterinary diagnostic labs are testing both human and animal samples. This exercise used blinded test samples and provided high confidence in the sensitivity of methods in twenty-nine laboratories for detection of SARS-CoV-2 variants while addressing the impact of sample matrix. Importantly, the results indicated that variants and matrix do not impact detection results. Additionally, this article examined decision-making criteria for Ct cut-off values from different laboratories and encouraged them to review and potentially reassess their criteria to improve future performance. This knowledge will lead to higher confidence in laboratory detection of current and new SARS-CoV-2 variants and aid in establishing reasonable cut-off parameters for these diagnostics tests.

## Introduction

COVID-19 illness caused by severe acute respiratory syndrome coronavirus 2 (SARS-CoV-2) has caused a worldwide pandemic in humans. The infection has also been reported in companion, farmed, zoo-managed, and wild animals around the world (1-10). In response to the spread of SARS-CoV-2 among many animal species, veterinary diagnostic laboratories are actively involved in SARS-CoV-2 detection in animal specimens. Laboratories also play a key role in the testing of millions of human samples for SARS-CoV-2. Among the available diagnostic tests, detection of the viral RNA with reverse transcription-quantitative polymerase chain reaction (RT-qPCR) is considered as the most widely used, rapid, and specific for diagnosis of the infection. To qualitatively and quantitatively evaluate the RT-qPCR methods routinely used for testing clinical specimens in veterinary diagnostic laboratories, the U. S. Food and Drug Administration (FDA) worked collaboratively with multiple organizations to offer two rounds of Interlaboratory Comparison (ILC) exercises in 2020 and 2021 (11, 12). The participants of the ILC exercises were able to detect RNA spiked in PrimeStore molecular transport medium (MTM). The method sensitivity and specificity were evaluated.

As SARS-CoV-2 continues to evolve, newly emerging variants present a major public health concern both due to their transmissibility and potential for immune evasion. In addition, because these variants can have extremely variable sequences, there is also concern about laboratory ability to detect sequences that may differ significantly from those of earlier variants (13). In ILC2, all participating laboratories were able to detect the Alpha and Beta variants with their routine methods (12). Since that time, additional variants emerged. The Delta variant (B.1.617.2) was initially identified in December 2020 and gained global attention due to its increased transmissibility in humans (14). The Delta variant has also infected animals, as evidenced by being transmitted from a fully vaccinated human to a canine in the United States in July 2021, and was later detected in white-tailed deer in Canada (5, 15). Additional animal species infected by the Delta variant include domestic cats and dogs, hamsters, lions, minks, gorillas, otters, snow leopards, ferrets, and hippopotamuses (14-20). Following the Delta emergence, a distantly related and even more heavily mutated and highly infectious variant Omicron (B.1.1.529) was identified in November 2021 and subsequently caused millions of illnesses in humans (16, 17). Phylogenetic analysis and gene sequencing of Omicron indicates potential zoonosis-associated mutations and host-jumping (21, 22). This variant has been found in mice, hamsters, and white-tailed deer (21, 23). Concerns of spillover of multiple lineages from animals to humans led to depopulation of farmed mink and pet hamsters (24, 25). To ensure reliable detection of SARS-CoV-2 in potentially infected animals and humans, it is critical for veterinary diagnostic laboratories to understand if their routine methods can detect current variants. This helps laboratories to consistently monitor their assays against emerging variants.

Nasal swab specimens are the primary matrix used for collecting respiratory samples, including those used for SARS-CoV-2 testing. In the previous ILC exercises (11, 12), MTM was used as matrix and diluent for the SARS-CoV-2 RNA panel preparation. In the current ILC3, nasal swabs were collected from dogs and used as a matrix for sample spiking. The use of canine nasal swab matrix allows for samples that are more reflective of real-life testing and may introduce additional factors into the samples that could complicate SARS-CoV-2 testing sensitivity.

When compared with the previous ILCs, the current ILC had the novelty of using inactivated virus instead of extracted RNA and using canine nasal matrix to replace MTM in the ILC samples. The overall goal of the study was to comparatively evaluate performance of veterinary diagnostic laboratories using blinded test samples. The specific objectives for ILC3 were (1) to explore the ability of ILC participants, using their routine RT-qPCR methods, to detect the presence of SARS-CoV-2 variants; (2) to evaluate the impact of canine nasal matrix on testing; and (3) to evaluate specificity of participants’ methods by including samples containing RNA from a non-SARS-CoV-2 animal coronavirus. The study was collaboratively conducted by: (i) the FDA Center for Veterinary Medicine’s Veterinary Laboratory Investigation and Response Network (Vet-LIRN), (ii) the Moffett Proficiency Testing (PT) Laboratory located at the Institute for Food Safety and Health at the Illinois Institute of Technology (IIT/IFSH) and the FDA Division of Food Processing Science and Technology, (iii) Kansas State University (KSU), (iv) Cornell University, (v) the United States Geological Survey (USGS), (vi) QuoData Quality and Statistics GmbH in Germany, (vii) US Department of Agriculture (USDA)’s National Veterinary Services Laboratories (NVSL) and National Animal Health Laboratory Network (NAHLN), and (viii) 29 participating laboratories.

## Materials and Methods

### Delta and Omicron Variants

Heat-inactivated SARS-CoV-2 Delta variant (B.1.617.2, Isolate hCoV-19/USA/MD-HP05285/2021, BEI Resources/ATCC NR-56128) and Omicron variant (B.1.1.529, hCoV-19/USA/GA-EHC-2811C/2021, BEI Resources/ATCC NR-56495) were purchased from BEI Resources (ATCC). According to the product information sheet, the virus was propagated in culture of *Homo sapiens* lung adenocarcinoma epithelial cells (Calu-3; ATCC® HTB-55™) infected with the isolate, and subsequently inactivated by heating to 65°C for 30 minutes. The Delta and Omicron concentrations were 8.62 × 10^5^ and 1.19 × 10^6^ genome equivalents per μL, respectively, as reported in the product Certificate of Analysis.

The concentration of received Delta variant was verified using droplet digital PCR (ddPCR) at Cornell University after RNA extraction performed at the ILC originating laboratory (FDA, Bedford Park, IL). Briefly, the virus was diluted in ten-fold serials in PrimeStore Molecular Transport Medium (MTM) (Longhorn) and its genomic RNA was extracted using the QIAamp Viral RNA Mini kit (Qiagen). The concentration of the extracted RNA was first estimated by comparing its RT-qPCR standard curve with that of quantitative synthetic SARS-CoV-2 RNA: ORF 1ab, E, N (ATCC VR-3276SD) of known target quantity (a serial dilution from 10 to 10^6^ copies per PCR reaction), on the Applied Biosystems 7500 Fast Real-Time PCR instrument (Thermo Fisher) with v.2.3 software, according to the Centers for Disease Control and Prevention (CDC) 2019-nCoV EUA kit method (26). The extracted RNA samples were then shipped to Cornell University for quantification on ddPCR (11, 12). The ddPCR was performed at the Cornell University Genomics Facility using the QX200 instrument (Bio-Rad), which used limiting dilutions of the target in up to 20,000 sub-nanoliter droplets to perform quantification without the use of a standard curve. The concentration of the Delta virus stock solution was determined as 1.5 × 10^6^ copies per μL. This concentration value was used for calculation during the ILC sample preparation.

### FIPV RNA Preparation

FIPV RNA was prepared at Cornell University as previously described (12). Briefly, the RNA was extracted from Fcwf-4 CU cell culture using the MagMAX™ Viral/Pathogen kit (Thermo Fisher). Quantification by ddPCR was performed as described above but with the P009 and P010 primers and P9/ P10 probe (27), which targets the *N* gene of FIPV.

### Nasal Matrix Preparation and Testing

Canine nasal swabs were collected, following the KSU Institutional Animal Care and Use Committee approved protocol #4318.1, from each nasal cavity of 12 healthy dogs housed at the KSU College of Veterinary Medicine for four rounds to accumulate enough matrix for the ILC exercise. Each swab was stored in 2 mL MTM and tested for SARS-CoV-2 using RT-qPCR and for canine respiratory pathogens according to the canine respiratory panel (https://vetview2.vet.k-state.edu/LabPortal/catalog/show/26303) available at KSU Veterinary Diagnostic Laboratory. Only swab samples that tested negative for SARS-CoV-2 and canine respiratory coronavirus were pooled and used for the exercise.

### Homogeneity and Stability of Delta Samples

To ensure ILC samples were appropriate for testing, acceptable homogeneity and stability were verified by the ILC originating Laboratory in two studies. In the first study, for each of the two analysts, five sets of 12 samples (S1 to S12) were prepared by adding the Delta variant to the nasal matrix at levels of 0, 3.75, 7.5, 37.5, 75, 750 or 7,500 copies per 100 µL (Supplemental Data, Table S1). The trial samples were inoculated and stored in the same manner as the ILC3 shipment samples. The trial sample sets were tested by Analyst A on Day 0, 7, 15, 21 and 43, and by Analyst B on Day 0, 7, 14, 21 and 28 after storage at -80 °C.

The second study was to analyze two sets of randomly chosen ILC samples prepared for shipment to participants: the 1st set was analyzed prior to the shipment day (pre-shipment, Table S4); and the 2nd set was analyzed one day after the shipment day (post-shipment, Table S5), to confirm the ILC samples were stable during the period from packing to arriving at the laboratories.

For analyzing the samples in the two studies, viral RNA was isolated using the QIAamp Viral RNA Mini Kit. The purified RNA was reverse transcribed to cDNA and amplified with specific primers and probes targeting two regions of the SARS-CoV-2 nucleocapsid gene (N1 and N2 markers, IDT), using AgPath-ID One-Step RT-PCR kit (Thermo-Fisher) (26). The RT-qPCR was performed on the Applied Biosystems 7500 Fast Real-Time PCR Instrument with version 2.3 software.

### Stability of Omicron Samples

Due to unavailability of the Omicron variant until three months after the Delta variant was purchased, the ILC originating laboratory was not able to include Omicron in the above stability and homogeneity studies. The viral concentration and its stability in MTM were verified by comparing its RT-qPCR standard curve with that of quantitative synthetic SARS-CoV-2 RNA: ORF 1ab, E, N (ATCC VR-3276SD) of known target quantity (a serial dilution from 10 to 10^6^ copies per PCR reaction). Briefly, two sets of Omicron samples were ten-fold serially diluted in MTM and stored at -80 °C (see Supplemental Data Table S6). On Day 0 and Day 21, the viral RNA was extracted using QIAamp Viral RNA Mini kit (Qiagen) and RT-qPCR was performed using N1 and N2 markers, as described above. The results indicated that Omicron in MTM was stable at -80 °C for up to 21 days.

### ILC3 Sample Preparation

Blank samples, blank with FIPV samples, and the Delta variant samples were prepared in nasal matrix (Table 1). Aliquots of 75 µL for each were distributed into 1.5-mL screw-top microfuge tubes. The Omicron variant samples were inoculated in MTM and aliquoted in the same manner. All samples were stored at -80 °C before shipping. To confirm successful inoculation, a set of ILC3 samples was tested at the ILC originating Laboratory (i.e., part of the Stability and Homogeneity Study-2) after they were prepared. The extracted RNA from this pre-shipment set was shipped to Cornell University for the spiking level verification using ddPCR. The ddPCR was performed as described above.

**Table 1:**
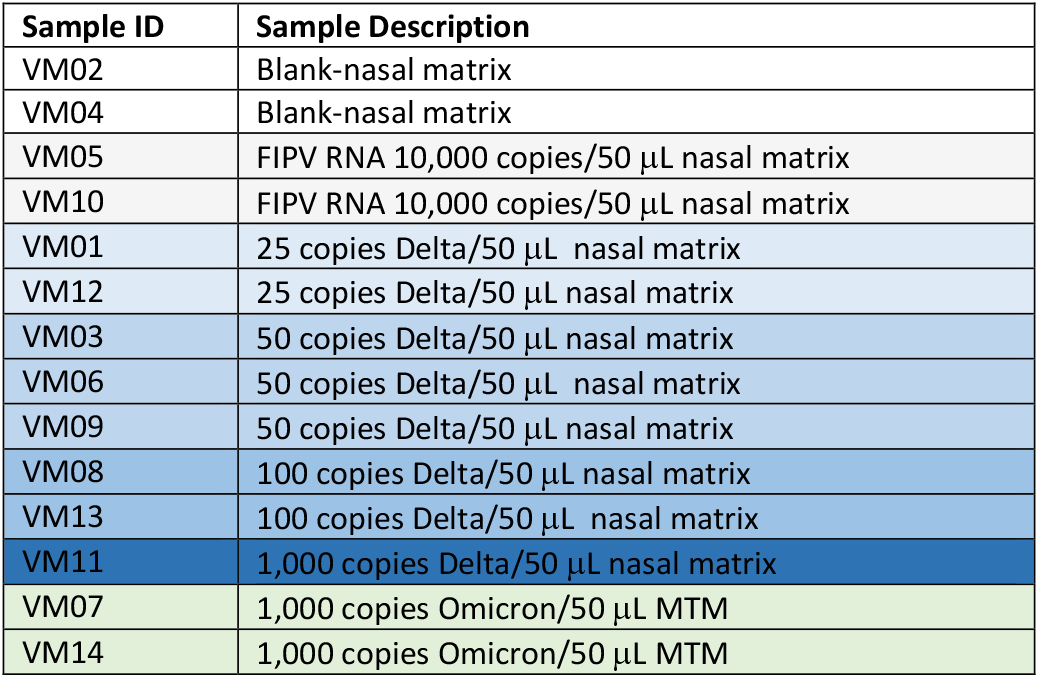
ILC3 sample composition.

| Sample ID | Sample Description |
| --- | --- |
| VM02 | Blank-nasal matrix |
| VM04 | Blank-nasal matrix |
| VM05 | FIPV RNA 10,000 copies/50 µL nasal matrix |
| VM10 | FIPV RNA 10,000 copies/50 µL nasal matrix |
| VM01 | 25 copies Delta/50 µL nasal matrix |
| VM12 | 25 copies Delta/50 µL nasal matrix |
| VM03 | 50 copies Delta/50 µL nasal matrix |
| VM06 | 50 copies Delta/50 µL nasal matrix |
| VM09 | 50 copies Delta/50 µL nasal matrix |
| VM08 | 100 copies Delta/50 µL nasal matrix |
| VM13 | 100 copies Delta/50 µL nasal matrix |
| VM11 | 1,000 copies Delta/50 µL nasal matrix |
| VM07 | 1,000 copies Omicron/50 µL MTM |
| VM14 | 1,000 copies Omicron/50 µL MTM |

### Sample Distribution and Result Submission

The final shipment samples were packaged in dry ice in a Saf-T-Pak STP-320 UN 3373 Category B Frozen Insulated Shipping System according to the manufacturer’s instructions and shipped via FedEx Priority Overnight.

Participants were instructed to take 50 μL from the 75 μL provided for testing by using a SARS-CoV-2 RNA detection method that they routinely use in their laboratories. After sample analysis, each participant submitted their results using an online portal. Results were reported as “Detected” (D), “Not Detected” (ND), or “Inconclusive” (IN) for SARS-CoV-2 viral RNA. The instructions required the participants to report Ct values, method information, and any modifications.

### Qualitative and Quantitative Assessment

Qualitative assessment was performed based on the rate of detection (ROD) values calculated separately for overall, N1 and N2 detection. For each of the three evaluations, ROD was calculated by dividing the number of “Detected” by the total number of test results at a particular copy number level. Inconclusive results were classified as “Not Detected”. The ROD for overall detection was based on the results reported in Table 2 and those for N1 and N2 detection were based on the Ct results shown in Table S7.

**Table 2:**
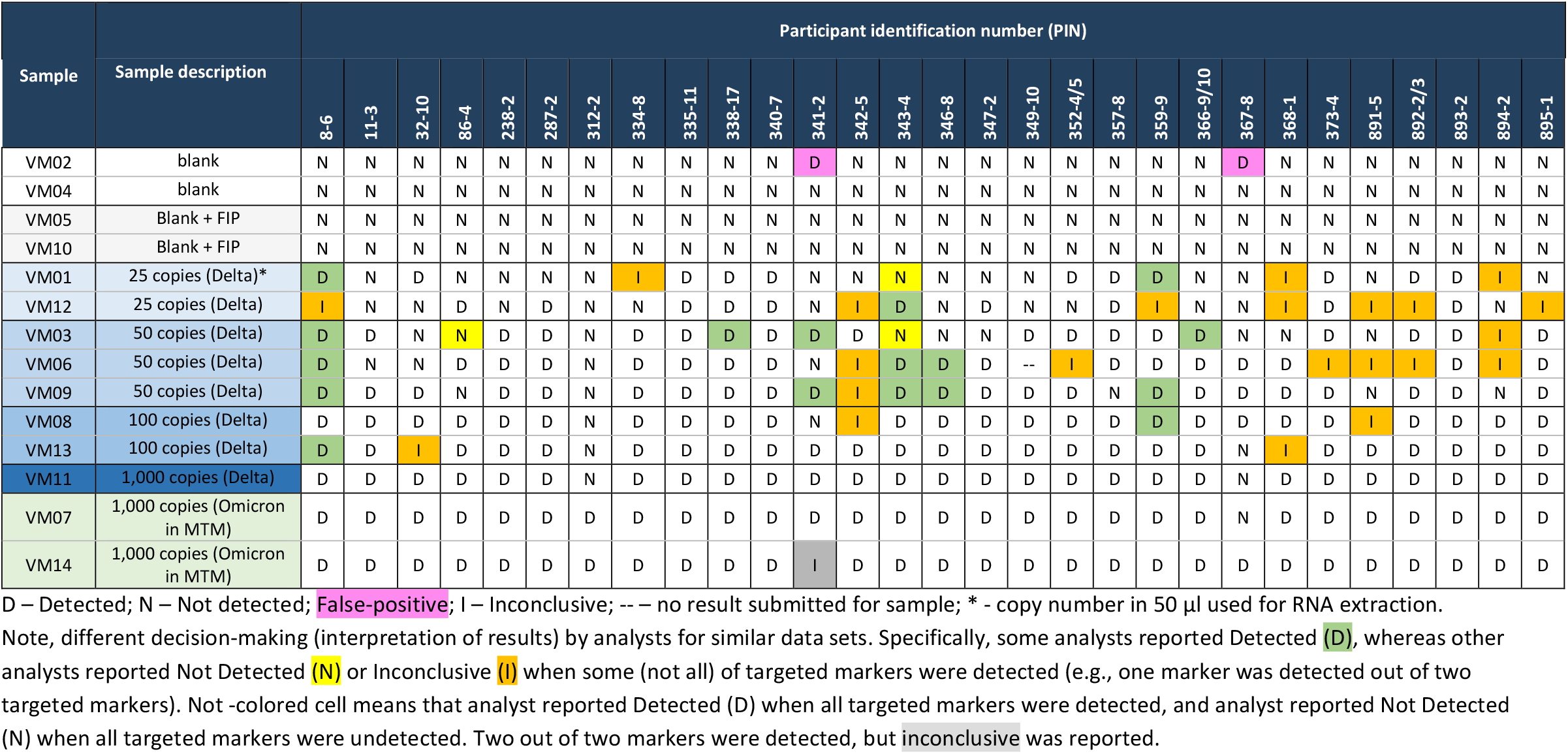
Results for “Overall Detection” submitted by the participants.

Quantitative assessment was performed based on mean values and standard deviations calculated using the Q/Hampel method (ISO 13528) (28). For participants who tested each sample in duplicate, triplicate or ran samples across multiple days, mean Ct values were used for statistical analysis. Repeatability standard deviation (s_r;ILC_), which characterizes the way that a parameter varies between the measurements performed by a single participant under near-identical testing conditions (on average across all participants), and reproducibility standard deviation (s_R;ILC_), which characterizes the way a given parameter varies between different participants, were calculated.

## Results

### Homogeneity and Stability Studies

Prior to ILC sample preparation, trials were conducted for homogeneity and stability analyses to ensure the same procedure to be applied for ILC sample preparation would produce homogeneous and stable samples. Study-1 results are shown in the Supplemental Data Table S1. Based on Study-1 results, sample stability was evaluated by calculated efficiency and level of detection at 95% probability of detection (LOD95) on different testing days (Table S2). For the evaluation of homogeneity, the sample standard deviation and the standard deviation for the duplicate measurements were determined (Table S3). These calculations were based exclusively on the Ct values for 3.125 copies/reaction (= 37.5 copies/100 µL) using the N1 marker. This level and marker combination yielded enough positive results for evaluation.

The homogeneity and stability Study-2 results are shown in the Supplemental Data Tables S4 and S5. A comparison of LOD95 and Ct values across Study-1 and Study-2 (Figure S1) showed that the pre- and post-shipment data were consistent with the stability data from Study-1.

The results demonstrated that the trial samples were deemed sufficiently homogenous and stable, and the inoculation process was suitable to prepare the designed ILC samples.

### Spiking Level Verification

The spiking levels in the ILC samples listed in Table 1 were confirmed using ddPCR. The extracted RNA from the pre-shipment set (i.e., the Study-2 sample set mentioned above) was shipped to Cornell University for ddPCR. Overall, the average range was within 40% of the theoretical value from extraction for both variants. These results were within expectations of the RNA extraction process.

### ILC Reported Results

Thirty participating laboratories were registered for this ILC, including Vet-LIRN, NAHLN, and private laboratories. Compared with ILC1 and ILC2, fewer laboratories were able to participate in ILC3 due to limitations of nasal matrix quantity. Thirty (30) data sets from 30 laboratories were received. One laboratory submitted late and requested to be excluded from statistical analysis.

The participants submitted qualitative “overall detection” results (Table 2) and Ct values for various markers (Tables S7-S10 in Supplemental Data). Effective volume from the original 50 µL sample was calculated based on the input and elution volumes for extraction and the RNA volume loaded into a PCR reaction that were reported by each participant (Table 3). For the 25 and 50 copies per 50 µL samples, 79% and 48% participants loaded less than 3.0 RNA copies to the PCR reaction, respectively. Such low copy numbers in the reaction often resulted in undetermined or high Ct results (e.g., Ct >37). However, eight participants (28%) were still able to detect the virus at the lowest level (25 copies per 50 μL in samples VM01 and VM012) with 1.4 or fewer copies per reaction (Table 2 and Table S7).

**Table 3:**
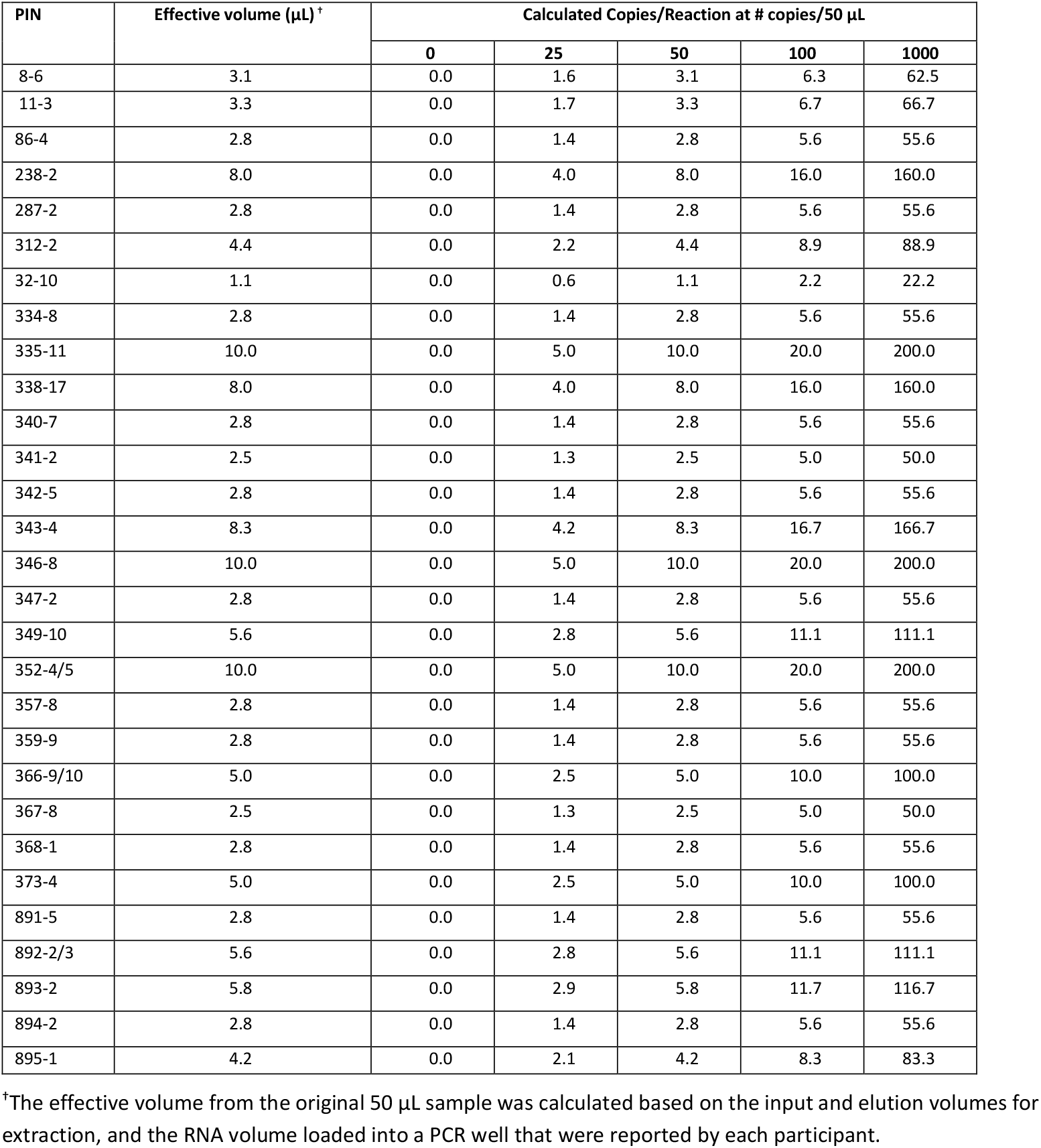
Effective volume and RNA copies per reaction used by the participants.

The “overall detection” results reported by participants (Table 2) were based on the Ct cut-off values established according to each laboratory’s internal RT-qPCR protocols (Table 4). The Ct cut-off values at 40, 44 or 45 were reported by 79% of ILC participants, often the last cycle number of the PCR, 17% participants set a Ct cut-off value at 37, one participant set it to 36, and one participant did not report a Ct cut-off value.

**Table 4:**
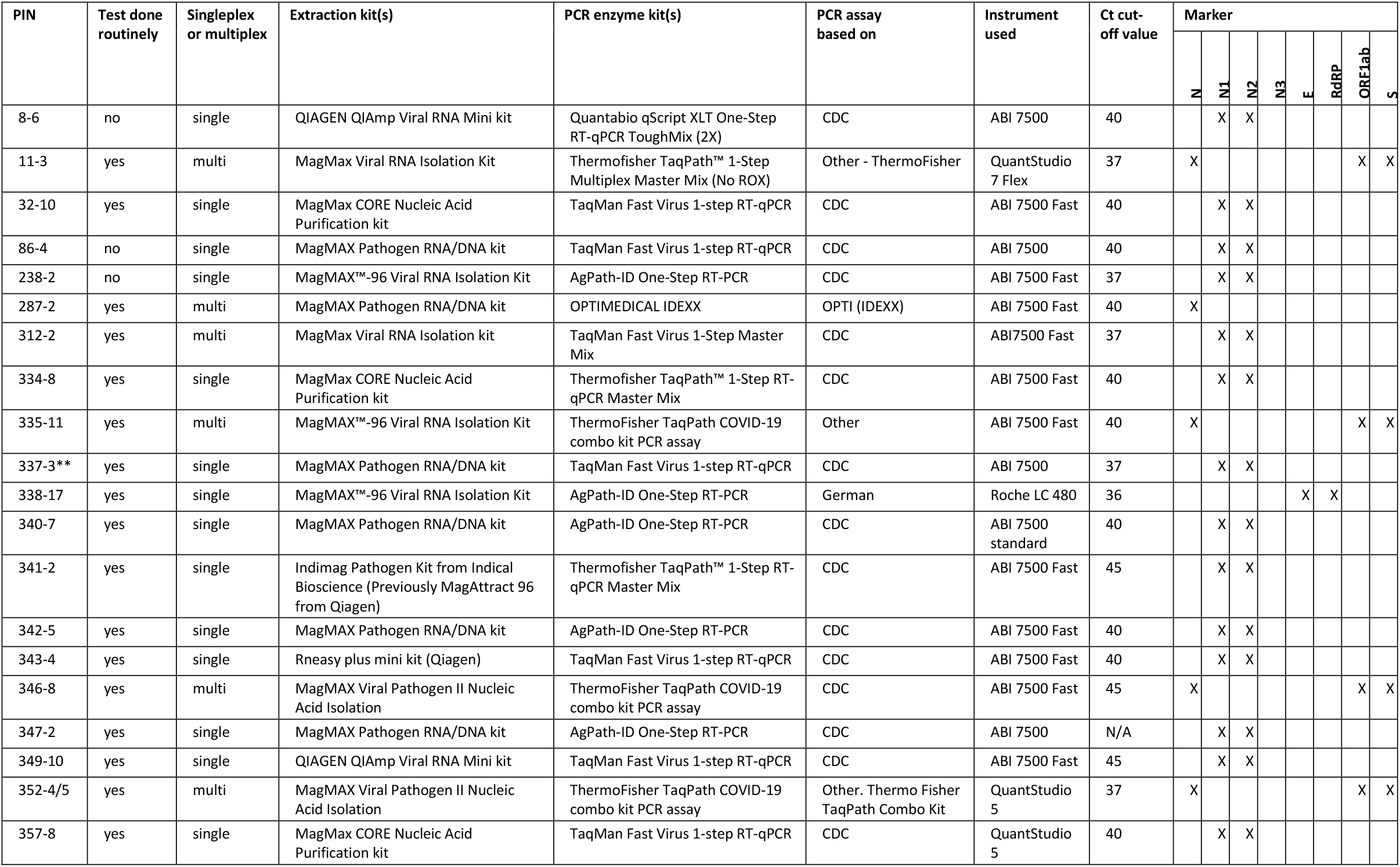
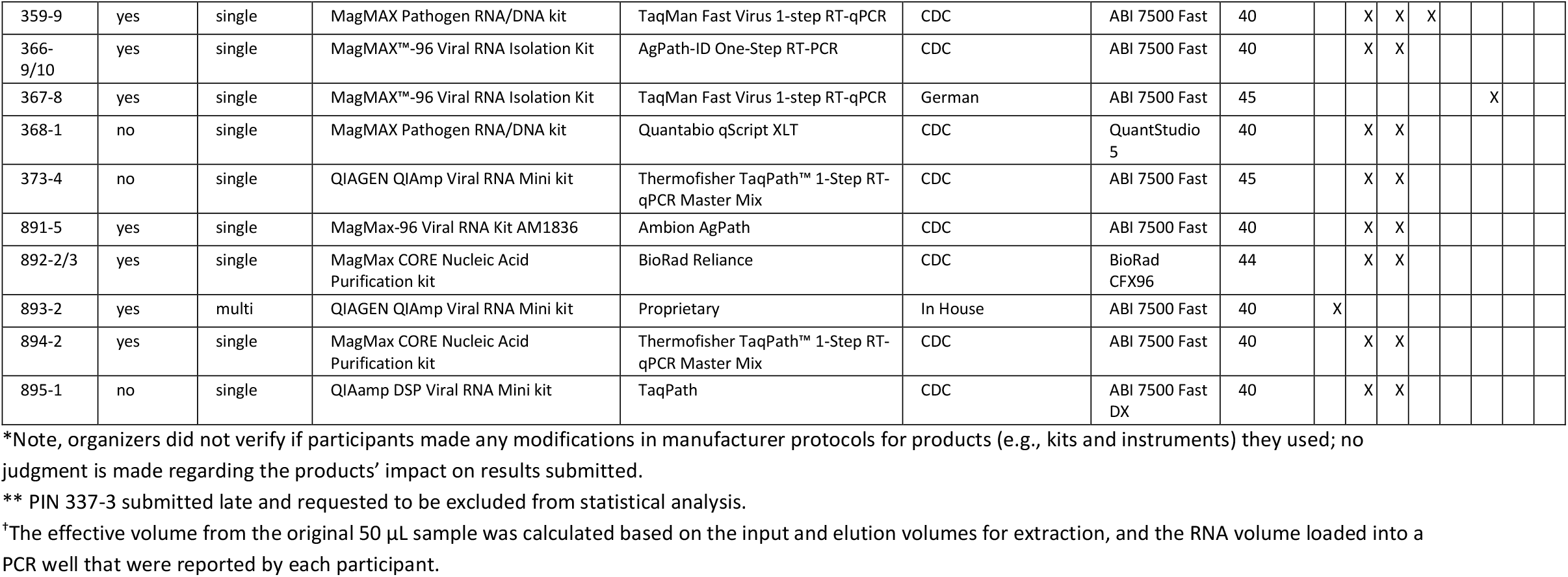
Overview of the method details and markers provided by the participants*.

Twenty-one participants submitted Ct values identified as corresponding to N1 and N2 markers (Table S7). Six participants submitted Ct values identified with N and one participant with N3 marker (Table S8). One participant submitted Ct values identified with E and two with RdRP marker (Table S9). Four participants submitted Ct values identified with ORFab1 and four with S marker (Table S10).

A total of 21 participants submitted Ct values obtained with internal controls, with 13 participants using Xeno (Thermo Fisher), five using MS2, and one each using beta actin QuantiNova (Qiagen) and the internal control provided by TetraCore. The internal control for all reporting participants performed as expected (data not shown). Specific method details and marker(s) used by participating laboratories are summarized in Table 4. A total of seven different nucleic acid extraction platforms and eight different PCR kits were used, including automated platforms and manual methods. Singleplex PCR was performed by 76% of the participants. The most frequently used PCR assay was based on the CDC methods, as observed for the previous two ILC exercises. Seventy-nine percent of the PCR assays were run on an ABI 7500 system, and 79% of the participants performed the SARS-CoV-2 test routinely. Participants were allowed to re-run a PCR assay if they needed to confirm an “inconclusive” result.

### Qualitative Assessment

For each participant at each spiking level, ROD values for N1, N2 and overall detection were calculated for qualitative assessment (Table 5).

**Table 5:** ROD (rate of detection) for overall, N1 and N2 detection.

| Level | Number of samples with same composition | Rate of detection (ROD) across participants |  |  |
| --- | --- | --- | --- | --- |
|  |  | Overall detection** | N1* | N2* |
| Number of laboratories included in the calculations |  | 29 | 21 | 21 |
| Blank-nasal matrix | 2 | 3 % | 2 % | 0 % |
| FIPV RNA 10,000 copies/50 µL nasal matrix | 2 | 0 % | 0 % | 0 % |
| 25 copies Delta (B.1.617.2) /50 µL nasal matrix | 2 | 36 % | 31 % | 38 % |
| 50 copies Delta (B.1.617.2) /50 µL nasal matrix | 3 | 70 % | 60 % | 56 % |
| 100 copies Delta (B.1.617.2) /50 µL nasal matrix | 2 | 86 % | 86 % | 83 % |
| 1,000 copies Delta (B.1.617.2) /50 µL nasal matrix | 1 | 93 % | 95 % | 95 % |
| 1,000 copies Omicron (B.1.1.52) /50 µL MTM | 2 | 97 % | 100 % | 100 % |

\* Ct values equal to or larger than the lab-specific cut-off are classified as “not detected”.
\*\* Inconclusive results classified as “not detected”.

“Overall detection” (i.e., the participants’ interpretation of their overall results) is shown in Table 2. Regarding the blank nasal matrix samples, a specificity of less than 100% was observed. The ROD was 3%, as two false positive results were submitted for one of the blank samples (VM02). Specifically, for the N1 marker, one false positive result was submitted for one of the blank samples (VM02), resulting in less than 100% specificity (i.e., ROD = 2%). The reported Ct values for false positive results were 40 for N1 reported by PIN 341-2 and 42 for RdRP reported by PIN 367-8. All the blank samples with FIPV RNA as confounder tested negative, indicating that the specificity of the routinely used methods was high.

For the Delta variant in nasal matrix samples, the ROD values increased with increasing copy numbers, consistently achieving ROD values above 93% at 1,000 copies per 50 µL for Delta variant for the overall, N1 and N2 evaluations. The ROD results of less than 100% at 1,000 copies per 50 µL were because two participants submitted negative results (false negative) for the Delta sample at 1,000 copies per 50 µL (VM11): (1) one participant (PIN 312-2) reported an overall negative result despite N1 Ct value meeting the Ct cutoff of 37, because N2 had a Ct of 38 (Table S7); (2) and the other participant (PIN 367-8) used only the RdRP marker for testing, with a negative result (Table S9).

For the Omicron variant in MTM samples, the ROD value for overall detection was also less than 100% at 1,000 copies. The reasons were: (1) the participant (PIN 367-8) who used only the RdRP marker submitted a negative for VM07 (Table S9); and (2) another participant (PIN341-2) reported an inconclusive (IN) result for VM14 even though the Ct values for both N1 and N2 were 35 and 40 which were actually lower than their cut-off value of 45 (Table S7). Based on the reported Ct values for N1 and N2, the ROD calculated for the two markers were 100% for the Omicron samples VM07 and VM14 (Table 5).

The Omicron samples (VM07 and VM14) were detected with all markers except the S marker (Table S10), likely due to multiple mutations in the *S* gene relative to earlier variants. Notably, all laboratories using the *S* gene had additional markers as part of their SARS-CoV-2 testing.

There was no significant difference in ROD values between the N1 and N2 marker for all samples.

### Quantitative Assessment

Mean values and standard deviations for the N1 and N2 markers were calculated according to the Q/Hampel method based on the submitted Ct values (Tables S7-S10) for quantitative assessment (Table 6). For the N1 and N2 markers, the mean Ct values decreased steadily with increasing spiking level, except for the mean Ct value at 25 copies per 50 µL being 0.36 lower than that at 50 copies per 50 µL for the N2 marker.

**Table 6:**
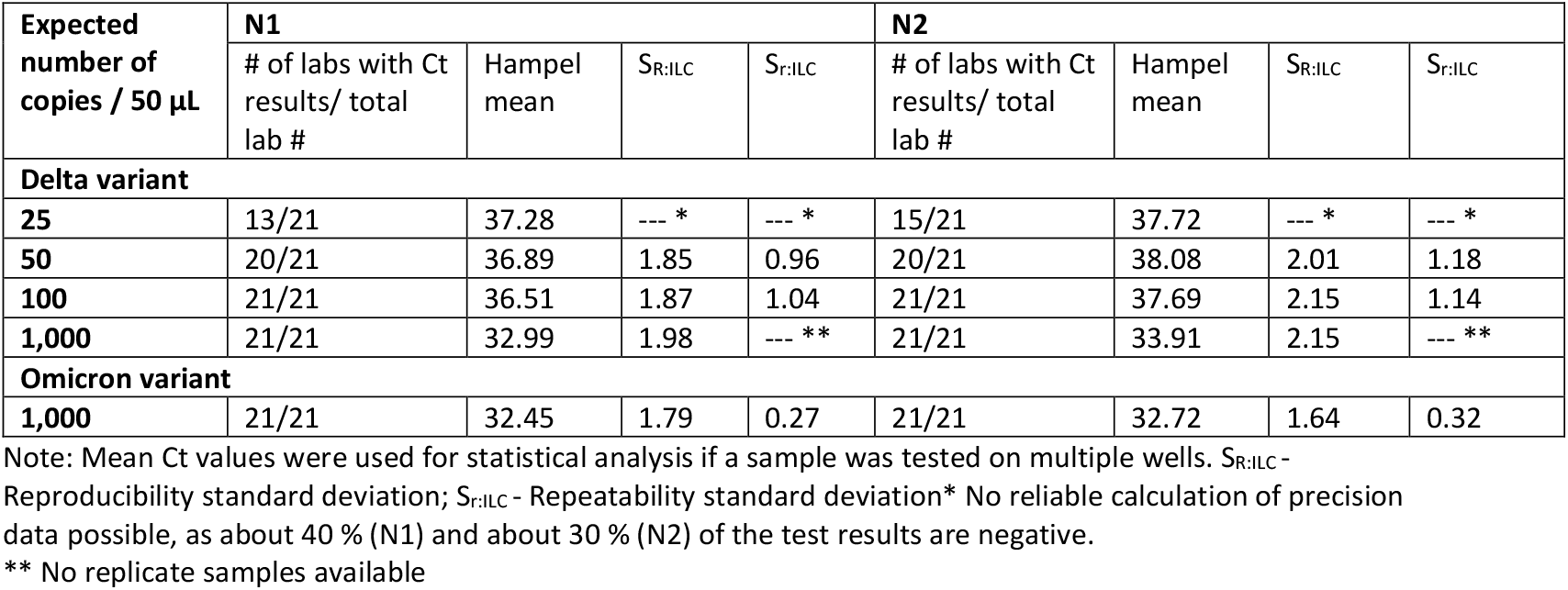
Statistical parameters for Ct values for N1 and N2 markers.

No significant differences were observed for samples containing comparable numbers of copies: neither between N1 and N2, nor between the Delta and the Omicron variants. Only for the Delta variant, Ct values seem to be slightly lower (about 1 Ct) for the N1 marker than for the N2 marker at the levels of 50 to 1,000 copies per 50 µL (Table 6).

The resulting reproducibility and repeatability standard deviations showed that no significant differences are among the variants, the copy numbers or the markers. On average, the reproducibility standard deviation (s_R;ILC_) between participants is slightly below 2 Ct for N1 and slightly higher than 2 Ct for N2. The repeatability standard deviation (s_r;ILC_) between samples in a single lab is around 1 Ct for both N1 and N2, again being slightly higher for N2.

## Discussion

The qualitative and quantitative assessment of the ILC3 data indicated that all the participants were able to detect Delta variant in canine nasal matrix and Omicron variant in MTM with overall sensitivity and specificity above 90%. However, several technical and strategic issues have been identified from the ILC and are discussed here.

To fulfill the ILC3 purpose of exploring testing ability, the participating laboratories were instructed to follow their routine procedures to perform the test, which led to interpretation of Ct results with a cut-off value established according to each laboratory’s internal RT-qPCR protocols as shown in Table 4, instead of applying a standardized Ct cut-off value across all participating laboratories. Comparing Ct values and cut-off values, the ILC organizers noted that individual participants used different decision-making criteria during interpretation of similar data sets. Specifically, some participants reported Detected (D), whereas others reported Not Detected (ND) or Inconclusive (I) when some (not all) of the targeted markers were detected (e.g., one marker was detected out of two targeted markers). This suggests an opportunity for laboratories to review and standardize their decision-making criteria during interpretation of Ct values when using multiple markers (i.e., to decide what to report when only one of two markers is detected). This ILC also suggests that the false negative rate and sensitivity of some methods can be improved if Ct cut-off values used are re-evaluated and optimized by analysts accordingly. It is especially important with Omicron, which often does not result in clinical signs in animals and presents at much lower virus levels in clinical samples when compared with Delta and earlier variants (29, 30).

It was also noted that false negative results were reported when a laboratory’s Ct cut-off value was set relatively low, such as Ct cut-off at 37. For many laboratories, the specimen tested is usually considered to be positive when a Ct value is obtained during the PCR run (e.g., a 40-cycle PCR). However, some laboratories would set a lower Ct cut-off value and consider the specimen as negative (i.e., false-negative) for any Ct values above the cut-off, by assuming the high Ct could be attributed to non-specific amplification (31).

Other than Ct cut-off value being the reason of false positive or false negative results, using a single marker for testing was another reason observed in the current ILC exercise. One laboratory that tested only the *RdRP* gene yielded multiple false negative and false positive results. *RdRP* was one of the commonly targeted genes used in Germany and France in 2020 (32). In a SARS-CoV-2 PCR kit comparison study (33), the authors reported that two kits targeting the *RdRP* gene [KAIRA kit (*E* and *RdRP* gene target) and PowerCheck kit (*RdRP* gene target)] yielded lower Ct values compared to others, which may explain why some laboratories rely on this marker exclusively. We conclude here that targeting multiple genes for SARS-CoV-2 PCR results in more reliable data than using a single marker. Providing this feedback to laboratories using single-target assays highlights the benefit of participating in an ILC and offers the laboratories valuable information to evaluate their technical procedures.

The Omicron variant carries a significant number of mutations on the *S* gene compared with other variants (34). Strategically, the participants who used the S marker for the ILC3 sample testing also used the N and ORFab1 markers which were able to detect the Omicron samples while the S marker missed them. The U.S. FDA identified molecular test kits that failed to detect Omicron and its sub-variants, and also recommended other kits with multiple gene targets for variants with *S*-gene or *N*-gene drop-out (13).

Components of the canine nasal cavity have the potential to impact viral detection assays. Additionally, the nasal cavity is filled with a species-rich bacterial community. Therefore, ILC3 used nasal matrix to better reflect actual testing conditions and determine potential effects on detection. Overall, specificity for the current ILC3 was 97%, as only 2 laboratories had false positives in blank samples containing nasal matrix. This was slightly less than that observed for ILC2 where specificity was over 99% for samples in MTM (12). Also, the variants tested changed from Alpha and Beta in ILC2 to Omicron and Delta in ILC3. Due to this, we cannot explicitly say that matrix and variants do not impact detection. However, none of the FIPV samples in canine nasal matrix resulted in SARS-CoV-2 positives. This suggests that the false positives in blank samples did not result from the change in matrix. These results along with the high specificity and sensitivity seen in ILC3 give confidence that matrix and variants did not impact assay performance.

The ROD values for ILC3 were similar to those for the ILC2 samples with similar spiking levels. For example, the ROD was 70% in this exercise versus 65% in ILC2 at 50 copies per 50 µL, and was 86% here compared to 85% in ILC2 at 100 copies per 50 µL. This indicates that neither the nasal matrix nor presence of different variants substantially affected overall SARS-CoV-2 detection. Importantly, the ROD (>80%) at the spiking level of 100 copies per 50 µL, where there may be only 5-20 copies per reaction in the RT-qPCR, indicated that the assays used by the participants are likely to work in diagnostic samples with high sensitivity.

## Conclusion

Overall, all ILC3 participating laboratories were able to detect both Delta and Omicron variants in canine nasal matrices or MTM. No significant impact on the detection of SARS-CoV-2 in the presence of the nasal matrix was observed. The RT-qPCR assays currently in use at the participating laboratories are able to detect a variety of variants, including those previously and currently circulating. In contrast to internal method validation, this ILC3 allowed participants to receive and analyze test samples of unknown composition within their own laboratory. Such evaluation provides a high degree of confidence that the assays used in veterinary diagnostic laboratories perform as expected during their routine testing of diagnostic samples. This study demonstrates the importance of ILCs to ensure confidence in laboratories performing SARS-CoV-2 testing of humans and animals to improve their performance as part of a comprehensive One Health pandemic response.

## Data Availability

All data produced in the present work are contained in the manuscript

## Non-standard Abbreviations

ILC: Interlaboratory comparison
RT–qPCR: quantitative reverse transcriptase–PCR or real-time reverse transcriptase–PCR
MTM: molecular transport medium
Ct: threshold cycle
ROD: rate of detection
LOD: level of detection
s.d.: standard deviation
FIPV: Feline infectious peritonitis virus.

## Acknowledgments

We acknowledge the diligence and hard work of the laboratory scientists who rapidly developed SARS-CoV-2 assays for animals and participated in this collaborative study. We acknowledge the following individuals for technical assistance and administrative support: Angelica Jones, Olgica Ceric, Ellen Hart, and Dave Rotstein from FDA, Joseph Flint from Cornell University, and Mia Torchetti and Christina Loiacono from USDA. We also thank Kirsten Simon from QuoData for supporting and facilitating this work. The Genomics Facility of the Biotechnology Resource Center at the Cornell University Institute of Biotechnology assisted with copy number quantification, and we thank Peter Schweitzer for facilitating this. We also thank Robert Newkirk from the FDA Proficiency Test and Method Validation Team for sample shipment assistance. We would like to acknowledge Steve Ensley, Lance Noll, Jianfa Bai, Rachel Palinski, and Paige Hess from Kansas State University for their work to collect and test the canine nasal matrix needed for this study. We would like to thank Andrew Scott and the Integrated Consortium of Laboratory Networks (ICLN) for their support and participation.

## Disclaimer

The views expressed in this article are those of the authors and do not necessarily reflect the official policy of the Department of Health and Human Services, the U.S. Food and Drug Administration, but do represent the views of the U.S. Geological Survey. The use of trade, firm, or product names is for descriptive purposes only and does not imply endorsement by the U.S. Government.

## Supplemental Data

Supplemental Data for this article is available online.

## References

1. USDA. 2020. Cases of SARS-CoV-2 in Animals in the United States. U.S. Department of Agriculture website: https://www.aphis.usda.gov/animal_health/one_health/downloads/sars-cov2-in-animals.pdf.

2. Sit TH, Brackman CJ, Ip SM, Tam KW, Law PY, To EM, Veronica Y, et al. Infection of dogs with SARS-CoV-2. Nature 2020;586:776–8.

3. Garigliany M, Van Laere AS, Clercx C, Giet D, Escriou N, Huon C, van der Werf S, et al. SARS-CoV-2 Natural Transmission from Human to Cat, Belgium, March 2020. Emerg Infect Dis 2020 Dec;26 12:3069–71. Epub 2020/08/14 as doi: 10.3201/eid2612.202223.

4. Jairak W, Charoenkul K, Chamsai E, Udom K, Chaiyawong S, Bunpapong N, Boonyapisitsopa S, et al. First cases of SARS-CoV-2 infection in dogs and cats in Thailand. Transbound Emerg Dis 2021 Nov 5 https://doi.org/10.1111/tbed.14383. Epub 2021/11/06 as doi: 10.1111/tbed.14383.

5. Wendling NM, Carpenter A, Liew A, Ghai RR, Gallardo-Romero N, Stoddard RA, Tao Y, et al. Transmission of SARS-CoV-2 Delta variant (B.1.617.2) from a fully vaccinated human to a canine in Georgia, July 2021. Zoonoses Public Health 2022 Apr 14. Epub 2022/04/16 as doi: 10.1111/zph.12944.

6. Oreshkova N, Molenaar RJ, Vreman S, Harders F, Oude Munnink BB, Hakze-van der Honing RW, Gerhards N, et al. SARS-CoV-2 infection in farmed minks, the Netherlands, April and May 2020. Euro Surveill 2020 Jun;25 23. Epub 2020/06/20 as doi: 10.2807/1560-7917.ES.2020.25.23.2001005.

7. McAloose D, Laverack M, Wang L, Killian ML, Caserta LC, Yuan F, Mitchell PK, et al. From people to panthera: natural SARS-CoV-2 infection in tigers and lions at the Bronx zoo. mBio 2020 Oct 13;11 5. Epub 2020/10/15 as doi: 10.1128/mBio.02220-20.

8. Hale VL, Dennis PM, McBride DS, Nolting JM, Madden C, Huey D, Ehrlich M, et al. SARS-CoV-2 infection in free-ranging white-tailed deer. Nature 2022 Feb;602 7897:481–6. Epub 2021/12/24 as doi: 10.1038/s41586-021-04353-x.

9. Kuchipudi SV, Surendran-Nair M, Ruden RM, Yon M, Nissly RH, Vandegrift KJ, Nelli RK, et al. Multiple spillovers from humans and onward transmission of SARS-CoV-2 in white-tailed deer. Proc Natl Acad Sci U S A 2022 Feb 8;119 6. Epub 2022/01/27 as doi: 10.1073/pnas.2121644119.

10. USDA. Confirmed cases of SARS-CoV-2 in animals in the United States. https://www.aphis.usda.gov/aphis/dashboards/tableau/sars-dashboard (Accessed May 16, 2022).

11. Deng K, Uhlig S, Ip HS, Killian ML, Goodman LB, Nemser S, Ulaszek J, et al. Interlaboratory comparison of SARS-CoV2 molecular detection assays in use by U.S. veterinary diagnostic laboratories. J Vet Diagn Invest 2021. Epub 2021 Julty 23.

12. Deng K, Uhlig S, Goodman LB, Ip HS, Killian ML, Nemser S, Ulaszek J, et al. Second round of the interlaboratory comparison (ILC) exercise of SARS-CoV-2 molecular detection assays being used by 45 veterinary diagnostic laboratories in the US. Journal of Veterinary Diagnostic Investigation 2022. Epub August 22, 2022 as doi: https://doi.org/10.1177/10406387221115702.

13. FDA. SARS-CoV-2 Viral Mutations: Impact on COVID-19 Tests. https://www.fda.gov/medical-devices/coronavirus-covid-19-and-medical-devices/sars-cov-2-viral-mutations-impact-covid-19-tests#detection-patterns (Accessed 2022 August 19).

14. Barroso-Arevalo S, Sanchez-Morales L, Perez-Sancho M, Dominguez L, Sanchez-Vizcaino JM. First Detection of SARS-CoV-2 B.1.617.2 (Delta) Variant of Concern in a Symptomatic Cat in Spain. Front Vet Sci 2022;9:841430. Epub 2022/04/19 as doi: 10.3389/fvets.2022.841430.

15. Kang K, Chen Q, Gao Y, Yu KJ. Detection of SARS-CoV-2 B.1.617.2 (Delta) variant in three cats owned by a confirmed COVID-19 patient in Harbin, China. Vet Med Sci 2022 May;8 3:945–6. Epub 2021/12/28 as doi: 10.1002/vms3.715.

16. Chan JF, Siu GK, Yuan S, Ip JD, Cai J, Chu AW, Chan W, et al. Probable Animal-to-Human Transmission of Severe Acute Respiratory Syndrome Coronavirus 2 (SARS-CoV-2) Delta Variant AY.127 Causing a Pet Shop-Related Coronavirus Disease 2019 (COVID-19) Outbreak in Hong Kong. Clinical Infectious Diseases 2022;ciac 171:1–6.

17. Doerksen T, Lu A, Noll L, Almes K, Bai J, Upchurch D, Palinski R. Near-Complete Genome of SARS-CoV-2 Delta (AY.3) Variant Identified in a Dog in Kansas, USA. Viruses 2021 Oct 19;13 10. Epub 2021/10/27 as doi: 10.3390/v13102104.

18. Jairak W, Chamsai E, Udom K, Charoenkul K, Chaiyawong S, Techakriengkrai N, Tangwangvivat R, et al. SARS-CoV-2 delta variant infection in domestic dogs and cats, Thailand. Sci Rep 2022 May 19;12 1:8403. Epub 2022/05/20 as doi: 10.1038/s41598-022-12468-y.

19. Mishra A, Kumar N, Bhatia S, Aasdev A, Kanniappan S, Sekhar AT, Gopinadhan A, et al. SARS-CoV-2 Delta Variant among Asiatic Lions, India. Emerg Infect Dis 2021 Oct;27(10):2723–2725 2021;27 10:2723-5.

20. Koeppel KN, Mendes A, Strydom A, Rotherham L, Mulumba M, Venter M. SARS-CoV-2 Reverse Zoonoses to Pumas and Lions, South Africa. Viruses 2022 Jan 11;14 1. Epub 2022/01/23 as doi: 10.3390/v14010120.

21. Pickering B, Lung O, Maguire F, Kruczkiewicz P, Kotwa JD, Buchanan T, Gagnier M, et al. Highly divergent white-tailed deer SARS-CoV-2 with potential deer-to-human transmission. bioRxiv 2022.

22. Wei C, Shan KJ, Wang W, Zhang S, Huan Q, Qian W. Evidence for a mouse origin of the SARS-CoV-2 Omicron variant. J Genet Genomics 2021 Dec;48 12:1111–21. Epub 2021/12/27 as doi: 10.1016/j.jgg.2021.12.003.

23. Diamond M, Halfmann P, Maemura T, Iwatsuki-Horimoto K, Iida S, Kiso M, Scheaffer S, et al. The SARS-CoV-2 B.1.1.529 Omicron virus causes attenuated infection and disease in mice and hamsters. Res Sq 2021 Dec 29. Epub 2022/01/05 as doi: 10.21203/rs.3.rs-1211792/v1.

24. Frutos R, Devaux CA. Mass culling of minks to protect the COVID-19 vaccines: is it rational? Microbes and New Infections 2020;38:100816.

25. Pang J, Siu T. Hong Kong to cull 2,000 hamsters after COVID-19 outbreak. Reuters 2022 Mya 10, 2022.

26. CDC. CDC 2019-novel coronavirus (2019-nCoV) real-time RT-PCR diagnostic panel for emergency use only instructions for use. https://www.fda.gov/media/134922/download (Accessed October 27, 2021).

27. Dye C, Helps CR, Siddell SG. Evaluation of real-time RT-PCR for the quantification of FCoV shedding in the faeces of domestic cats. J Feline Med Surg 2008 Apr;10 2:167–74. Epub 2008/02/05 as doi: 10.1016/j.jfms.2007.10.010.

28. Uhlig S, Colson B, Gowik P. Taking laboratory uncertainties into account in the Hampel estimator. Accreditation and Quality Assurance 2018;24 as doi: 10.1007/s00769-018-1332-x.

29. Sanchez-Morales L, Sanchez-Vizcaino JM, Perez-Sancho M, Dominguez L, Barroso-Arevalo S. The Omicron (B.1.1.529) SARS-CoV-2 variant of concern also affects companion animals. Frontiers in Evterinary Science 2022 as doi: 10.3389/fvets.2022.940710.

30. Mathias M, Nascimento GM, Nooruzzaman M, Yuan F, Chen C, Caserta LC, Miller AD, et al. The Omicron variant BA.1.1 presents a lower pathogenicity than B.1 D614G and Delta variants in a feline model of SARS-CoV-2 infection. bioRxiv 2022 as doi: 10.1101/2022.06.15.496220.

31. Caraguel CG, Stryhn H, Gagne N, Dohoo IR, Hammell KL. Selection of a cutoff value for real-time polymerase chain reaction results to fit a diagnostic purpose: analytical and epidemiologic approaches. J Vet Diagn Invest 2011 Jan;23 1:2–15. Epub 2011/01/11 as doi: 10.1177/104063871102300102.

32. Barreto HG, de Padua Milagres FA, de Araujo GC, Daude MM, Benedito VA. Diagnosing the novel SARS-CoV-2 by quantitative RT-PCR: variations and opportunities. J Mol Med (Berl) 2020 Dec;98 12:1727–36. Epub 2020/10/18 as doi: 10.1007/s00109-020-01992-x.

33. Altamimi AM, Obeid DA, Alaifan TA, Taha MT, Alhothali MT, Alzahrani FA, Albarrag AM. Assessment of 12 qualitative RT-PCR commercial kits for the detection of SARS-CoV-2. J Med Virol 2021 May;93 5:3219–26. Epub 2021/02/26 as doi: 10.1002/jmv.26900.

34. Salleh MZ, Derrick JP, Deris ZZ. Structural Evaluation of the Spike Glycoprotein Variants on SARS-CoV-2 Transmission and Immune Evasion. Int J Mol Sci 2021 Jul 10;22 14. Epub 2021/07/25 as doi: 10.3390/ijms22147425.

