## Supplemental material for "Successful Detection of Delta and Omicron Variants of SARS-CoV-2 by Veterinary Diagnostic Laboratory Participants in an Interlaboratory Comparison Exercise"

Any use of trade, firm, or product names is for descriptive purposes only and does not imply endorsement by the U.S. Government.

### 1. Evaluation of homogeneity and stability in Study-1 and Study-2

#### 1.1 Stability and homogeneity Study-1

Two trials were conducted for Study-1. For each trial, five sets of 12 samples (SARS-CoV-2 Delta variant in nasal matrix) described below were inoculated and stored in the same manner as the ILC shipment samples. One set of samples was tested by Analyst A on Day 0, 7, 15, 21 and 43, and by Analyst B on Day 0, 7, 14, 21 and 28 after storage at -80 °C. All tests were performed on the same PCR instrument.

Sample concentration levels were as follows:

| Trial sample name | Delta variant level |
|---|---|
| S1-S2 | 0 copies per 100 μL |
| S3-S4 | 3.75 copies per 100 μL |
| S5-S6 | 7.5 copies per 100 μL |
| S7-S9 | 37.5 copies per 100 μL |
| S10 | 75 copies per 100 μL |
| S11 | 750 copies per 100 μL |
| S12 | 7,500 copies per 100 μL |

S: sample

**Table S1: Testing results for Study-1**

| Testing Day | Sample | Delta variant Inoculation Level (copies/100 μL) | Trial 1-Analyst A Results | Trial 2-Analyst B Results |
|---|---|---|---|---|
| 0 | S1 | 0 | ND | ND |
| | S2 | | ND | ND |
| | S3 | 3.75 | ND | ND |
| | S4 | | ND | ND |
| | S5 | 7.5 | D | ND |
| | S6 | | D | ND |
| | S7 | 37.5 | D | D |
| | S8 | | D | D |

| | | | | |
|---|---|---|---|---|
| | S9 | | D | D |
| | S10 | 75 | D | D |
| | S11 | 750 | D | D |
| | S12 | 7,500 | D | D |
| 7 | S1 | 0 | ND | ND |
| | S2 | | ND | ND |
| | S3 | 3.75 | ND | ND |
| | S4 | | ND | ND |
| | S5 | 7.5 | ND | D |
| | S6 | | D | ND |
| | S7 | 37.5 | D | D |
| | S8 | | D | D |
| | S9 | | D | D |
| | S10 | 75 | D | D |
| | S11 | 750 | D | D |
| | S12 | 7,500 | D | D |
| 14 | S1 | 0 | | ND |
| | S2 | | | ND |
| | S3 | 3.75 | | ND |
| | S4 | | | ND |
| | S5 | 7.5 | | D |
| | S6 | | | D |
| | S7 | 37.5 | | D |
| | S8 | | | D |
| | S9 | | | D |
| | S10 | 75 | | D |
| | S11 | 750 | | D |
| | S12 | 7,500 | | D |
| 15 | S1 | 0 | ND | |
| | S2 | | ND | |
| | S3 | 3.75 | D | |
| | S4 | | D | |
| | S5 | 7.5 | D | |
| | S6 | | D | |
| | S7 | 37.5 | D | |
| | S8 | | D | |
| | S9 | | D | |
| | S10 | 75 | D | |
| | S11 | 750 | D | |
| | S12 | 7,500 | D | |

| | | | | |
|---|---|---|---|---|
| 21 | S1 | 0 | ND | ND |
| | S2 | | ND | ND |
| | S3 | 3.75 | ND | D |
| | S4 | | ND | ND |
| | S5 | 7.5 | ND | D |
| | S6 | | D | ND |
| | S7 | 37.5 | D | D |
| | S8 | | D | D |
| | S9 | | D | D |
| | S10 | 75 | D | D |
| | S11 | 750 | D | D |
| | S12 | 7,500 | D | D |
| 28 | S1 | 0 | | ND |
| | S2 | | | ND |
| | S3 | 3.75 | | D |
| | S4 | | | D |
| | S5 | 7.5 | | ND |
| | S6 | | | ND |
| | S7 | 37.5 | | D |
| | S8 | | | D |
| | S9 | | | D |
| | S10 | 75 | | D |
| | S11 | 750 | | D |
| | S12 | 7,500 | | D |
| 43 | S1 | 0 | ND | |
| | S2 | | ND | |
| | S3 | 3.75 | ND | |
| | S4 | | ND | |
| | S5 | 7.5 | ND | |
| | S6 | | ND | |
| | S7 | 37.5 | D | |
| | S8 | | D | |
| | S9 | | D | |
| | S10 | 75 | D | |
| | S11 | 750 | D | |
| | S12 | 7,500 | D | |

Note: Each set of Study-1 samples was extracted by using Qiagen QIAamp Viral RNA Mini kit for manual RNA extraction. Briefly, the RNA was isolated from the 100 µL samples and 60 µL was eluted from the Qiagen kit column. The extracted RNA (5 µL) was analyzed using AgPath-ID One-Step RT-PCR kit. S: Sample, D: Determined by either N1 and/or N2 marker; ND: not determined by both N1 and N2 markers.

### 1.2 Evaluation of stability in Study-1

In Table S2, the results of the stability test are given. The last column represents the LOD95 in relation to effective volume, whereas the other columns represent results based on quantitative Ct values. The regression parameters were calculated on the basis of the $\log_{10}$ copy numbers per reaction. Note that new primers and probes were used on Day 28 for analyst B and this may be related to the improved performance for N2 over time for analyst B.

**Table S2: Results of stability testing**

| Analyst | Target | Day | Number of data points | Regression Parameters | | Efficiency [%] | Expected CT Values Number of Copies per Reaction | | | | | | LOD95 |
|---|---|---|---|---|---|---|---|---|---|---|---|---|---|
| | | | | Intercept | Slope | | 0.3125 | 0.625 | 3.125 | 6.25 | 62.5 | 625 | |
| | | 0 | 13 | 36.70 | -2.92 | 120 | 38.17 | 37.29 | 35.25 | 34.38 | 31.46 | 28.54 | 5 |
| | | 7 | 11 | 37.75 | -3.46 | 95 | 39.50 | 38.46 | 36.04 | 35.00 | 31.54 | 28.09 | 8 |
| | N1 | 15 | 16 | 36.26 | -2.71 | 134 | 37.63 | 36.82 | 34.92 | 34.11 | 31.40 | 28.69 | 18 |
| | | 21 | 12 | 37.44 | -3.41 | 97 | 39.16 | 38.14 | 35.76 | 34.73 | 31.33 | 27.92 | 6 |
| A | | 43 | 12 | 37.95 | -3.66 | 88 | 39.80 | 38.69 | 36.13 | 35.03 | 31.37 | 27.71 | 5 |
| | | 0 | 9 | 40.62 | -3.32 | 100 | 42.29 | 41.29 | 38.97 | 37.98 | 34.66 | 31.34 | 13 |
| | | 7 | 11 | 40.34 | -3.12 | 109 | 41.92 | 40.98 | 38.80 | 37.86 | 34.74 | 31.63 | 8 |
| | N2 | 15 | 11 | 40.39 | -3.18 | 106 | 41.99 | 41.04 | 38.81 | 37.86 | 34.68 | 31.50 | 8 |
| | | 21 | 10 | 40.90 | -3.53 | 92 | 42.69 | 41.63 | 39.15 | 38.09 | 34.56 | 31.02 | 10 |
| | | 43 | 12 | 40.48 | -3.59 | 90 | 42.29 | 41.21 | 38.70 | 37.62 | 34.03 | 30.45 | 5 |
| | | 0 | 11 | 36.71 | -3.03 | 114 | 38.24 | 37.33 | 35.21 | 34.30 | 31.28 | 28.25 | 7 |
| | | 7 | 13 | 36.66 | -3.02 | 115 | 38.18 | 37.27 | 35.17 | 34.26 | 31.24 | 28.23 | 5 |
| | N1 | 14 | 13 | 36.60 | -3.05 | 113 | 38.14 | 37.23 | 35.09 | 34.17 | 31.12 | 28.07 | 5 |
| | | 21 | 13 | 36.85 | -2.77 | 129 | 38.25 | 37.42 | 35.48 | 34.65 | 31.88 | 29.10 | 5 |
| B | | 28 | 12 | 38.14 | -3.79 | 84 | 40.05 | 38.91 | 36.26 | 35.12 | 31.33 | 27.54 | 5 |
| | | 0 | 6 | 42.07 | -4.02 | 77 | 44.10 | 42.89 | 40.08 | 38.87 | 34.85 | 30.83 | 40 |

| | | | | | | | | | | | | |
|---|---|---|---|---|---|---|---|---|---|---|---|---|
| | 7 | 9 | 40.98 | -3.68 | 87 | 42.84 | 41.73 | 39.16 | 38.05 | 34.38 | 30.70 | 15 |
| **N2** | 14 | 8 | 39.83 | -3.02 | 115 | 41.36 | 40.45 | 38.34 | 37.43 | 34.41 | 31.40 | 21 |
| | 21 | 6 | 41.27 | -3.23 | 104 | 42.90 | 41.92 | 39.67 | 38.69 | 35.46 | 32.23 | 45 |
| | 28 | 10 | 39.07 | -2.58 | 144 | 40.37 | 39.60 | 37.80 | 37.02 | 34.45 | 31.87 | 12 |

### 1.3 **Evaluation of homogeneity in Study-1**

Based on the Ct values for 3.125 copies/reaction, the standard deviation for the duplicate measurements ($s_e$), the sample standard deviation ($s_{sample}$) and the mean Ct values were calculated. The results for the N1 marker are given in Table S3 – separately for each level, analyst, and day. The evaluation for a combination of target/level/analyst/day was only possible if there were at least two samples with two replicates each. Only those samples were considered for which both determinations yielded positive results, i.e., Ct values were available. Since the majority of the results for N2 were negative for 3.125 copies/reaction, the evaluation of homogeneity could only be carried out for N1.

**Table S3: Summary of standard deviation for the duplicate measurements ($s_e$), sample standard deviation ($s_{sample}$), and mean Ct value**

| Analyst | Day | $s_e$ | $s_{sample}$ | Mean Ct value |
|---|---|---|---|---|
| **A** | 0 | 0.50 | 0.21 | 35.21 |
| | 7 | 0.67 | 0.22 | 36.31 |
| | 15 | 0.42 | 0.75 | 36.20 |
| | 21 | 1.07 | 0.00 | 36.18 |
| | 43 | 0.46 | 0.26 | 36.11 |
| | *Average* | *0.67* | *0.38* | *36.00* |
| **B** | 0 | 1.20 | 0.00 | 35.02 |
| | 7 | 0.20 | 0.00 | 35.65 |
| | 14 | 0.33 | 0.00 | 35.50 |
| | 21 | 0.91 | 0.00 | 36.33 |
| | 28 | 0.45 | 0.00 | 36.36 |
| | *Average* | *0.72* | *0.00* | *35.77* |

### 1.4 **Stability and homogeneity results for Study-2**

#### Table S4: Pre-shipment testing data

A set of samples was tested at the ILC originating laboratory prior to shipping. An aliquot of 50 μL from each sample was used for Qiagen QIAamp Viral RNA Mini kit extraction and 60 μL extracted RNA was eluted from the Qiagen column. The extracted RNA (5 μL) was analyzed using AgPath-ID One-Step RT-PCR kit with N1 and N2 markers.

| Sample | Inoculation Level | Results |
|---|---|---|
| VM01-04252022 | 25 copies Delta/50 mL nasal matrix | D |
| VM02-04252022 | Blank-nasal matrix | ND |
| VM03-04252022 | 50 copies Delta/50 mL nasal matrix | D |
| VM04-04252022 | Blank- nasal matrix | ND |
| VM05-04252022 | Blank- FIPV RNA 10,000 copies/50 mL nasal matrix | ND |
| VM06-04252022 | 50 copies Delta/50 mL nasal matrix | D |
| VM07-04252022 | 1000 copies Omicron/50 mL MTM | D |
| VM08-04252022 | 100 copies Delta/50 mL nasal matrix | D |
| VM09-04252022 | 50 copies Delta/50 mL nasal matrix | D |
| VM10-04252022 | Blank- FIPV RNA 10,000 copies/50 mL nasal matrix | ND |
| VM11-04252022 | 1,000 copies Delta/50 mL nasal matrix | D |
| VM12-04252022 | 25 copies Delta/50 mL nasal matrix | D |
| VM13-04252022 | 100 copies Delta/50 mL nasal matrix | D |
| VM14-04252022 | 1000 copies Omicron/50 mL MTM | D |

D: Determined by either N1 and/or N2 marker; ND: not determined by both N1 and N2 markers.

#### Table S5: Post-shipment testing data

A set of samples was tested at the ILC originating laboratory after shipping. Same procedures were performed as described for the pre-shipment test.

| Sample | Inoculation Level | Results |
|---|---|---|
| VM01-04252022 | 25 copies Delta/50 mL nasal matrix | D |
| VM02-04252022 | Blank-nasal matrix | ND |

| | | |
|---|---|---|
| VM03-04252022 | 50 copies Delta/50 mL nasal matrix | D |
| VM04-04252022 | Blank- nasal matrix | ND |
| VM05-04252022 | Blank- FIPV RNA 10,000 copies/50 mL nasal matrix | ND |
| VM06-04252022 | 50 copies Delta/50 mL nasal matrix | D |
| VM07-04252022 | 1000 copies Omicron/50 mL MTM | D |
| VM08-04252022 | 100 copies Delta/50 mL nasal matrix | D |
| VM09-04252022 | 50 copies Delta/50 mL nasal matrix | D |
| VM10-04252022 | Blank- FIPV RNA 10,000 copies/50 mL nasal matrix | ND |
| VM11-04252022 | 1,000 copies Delta/50 mL nasal matrix | D |
| VM12-04252022 | 25 copies Delta/50 mL nasal matrix | D |
| VM13-04252022 | 100 copies Delta/50 mL nasal matrix | D |
| VM14-04252022 | 1000 copies Omicron/50 mL MTM | D |

D: Determined by either N1 and/or N2 marker; ND: not determined by both N1 and N2 markers.

### 1.5 **Evaluation of stability across Study-1 and Study-2**

In Figure S1, a comparison of the LOD95 and Ct values for 3.125 copies/reaction (Ct values calculated based on regression analysis) of stability data (Study-1) as well as pre- and post-shipment data (Study-2) is given.

**Figure S1: Comparison of LOD95 and Ct values for 3.125 copies/reaction across Study-1 and Study-2**

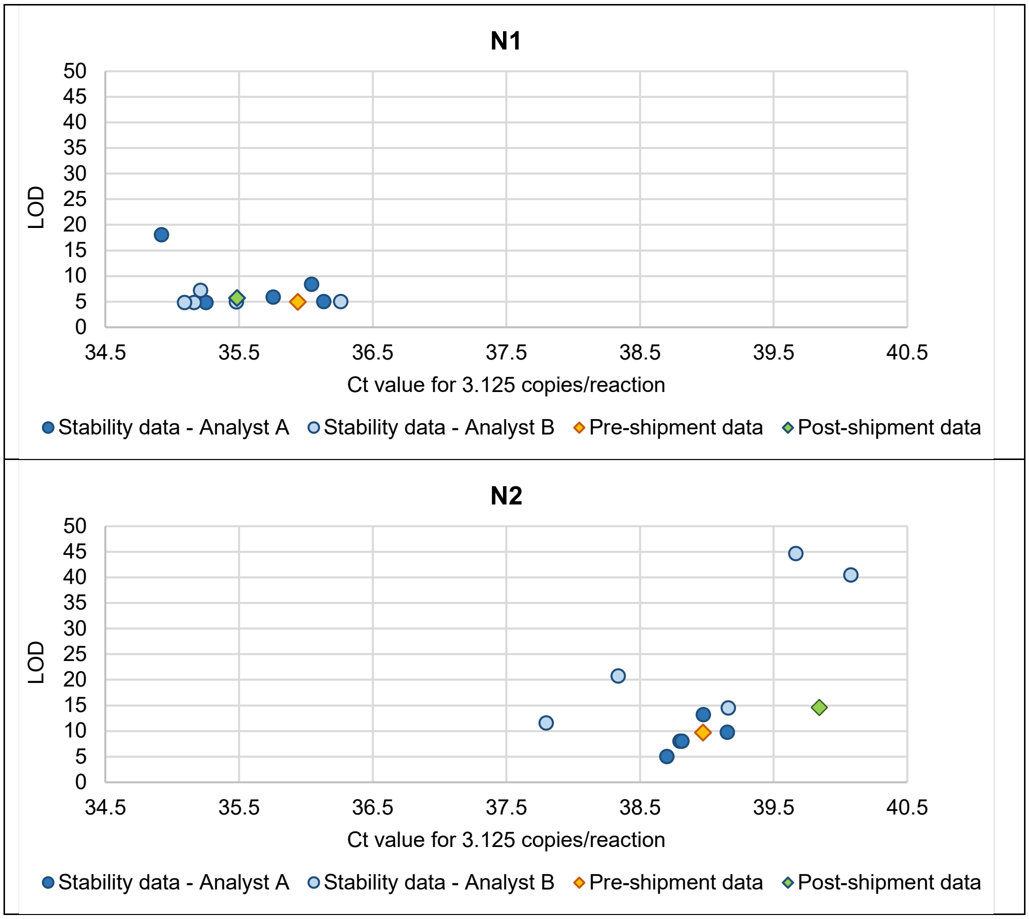

#### 1.6 **Stability of Omicron variant in MTM**

Two sets of ten-fold serial diluted SARS-CoV-2 Omicron variant in MTM were prepared and stored at -80 °C. On Day 0 and Day 21, the viral RNA was extracted using QIAamp Viral RNA Mini kit and RT-qPCR was performed using the N1 and N2 markers, as described above at the ILC originating laboratory.

**Table S6: Stability of Omicron in MTM**

| Testing Day | Sample | Omicron Variant Inoculation Level (copies/50 µL) | Results |
|---|---|---|---|
| 0 | S1 | 0 | ND |
| | S2 | 60 | D |
| | S3 | 600 | D |
| | S4 | 6,000 | D |
| | S5 | 60,000 | D |
| | S6 | 600,000 | D |
| 21 | S1 | 0 | ND |
| | S2 | 60 | D |
| | S3 | 600 | D |
| | S4 | 6,000 | D |
| | S5 | 60,000 | D |
| | S6 | 600,000 | D |

D: Determined by either N1 and/or N2 marker; ND: not determined by both N1 and N2 markers.

### 2. Ct values submitted by the participating laboratories

**Table S7: Ct values submitted by the laboratories for the N1 and N2 markers**

| Marker | Sample | Sample description | Participant Identification Number (PIN) | | | | | | | | | | | | | | | | | | | | | | | | | | | | | |
|---|---|---|---|---|---|---|---|---|---|---|---|---|---|---|---|---|---|---|---|---|---|---|---|---|---|---|---|---|---|---|---|---|
| | | | 8-6 | 11-3 | 32-10 | 86-4 | 238-2 | 287-2 | 312-2 | 334-8 | 335-11 | 337-3 | 338-17 | 340-7 | 341-2 | 342-5 | 343-4 | 346-8 | 347-2 | 349-10 | 352-4/5 | 357-8 | 359-9 | 366-9/10 | 367-8 | 368-1 | 373-4 | 891-5 | 892-2/3 | 893-2 | 894-2 | 895-1 |
| N1 | VM02 | blank | n/n | | n | n | n | | n | n | | n | | n | 40 | n | n/n | | n | n | | n | n | n | | n | n | n | n/n | | n | n |
| | VM04 | blank | n/n | | n | n | n | | n | n | | n | | n | n | n | n/n | | n | n | | n | n | n | | n | n | n | n/n | | n | n |
| | VM05 | Blank + FIP | n/n | | n | n | n | | n | n | | n | | n | n | n | n/n | | n | n | | n | n | n | | n | n | n | n/n | | n | n |
| | VM10 | Blank + FIP | n/n | | n | n | n | | n | n | | n | | n | n | n | n/n | | n | n | | n | n | n | | n | n | n | n/n | | n | n |
| | VM01 | 25 copies (Delta)* | 36/n | | 34 | 39 | n | | 39 | n | | n | | 35 | n | n | 38/n | | n | n | | 36 | n | n | | 40 | 36 | n | 39/37 | | 37 | n |
| | VM12 | 25 copies (Delta) | n/37 | | n | 39 | n | | n | n | | n | | 36 | n | n | 38/n | | 37 | n | | 38 | n | n | | n* | 36 | 37 | 39/n | | n | n |
| | VM03 | 50 copies (Delta) | n/37 | | n/n/n | n | 35 | | n | 34 | | 35 | | 35 | n | 37 | n | | n | 38 | | 36 | 34 | 39 | | n | 37 | n | 40/40 | | 36 | 33 |
| | VM06 | 50 copies (Delta) | 34/n | | n | 37 | 36 | | n | 35 | | n | | 36 | n | 37 | 38/n | | 38 | --- | | 37 | 36 | 38 | | 38 | 36 | n | 39/n | | 37 | 34 |
| | VM09 | 50 copies (Delta) | 35/n | | 36 | n | 36 | | 38 | 35 | | 35 | | 37 | 39 | n | 39/38 | | 36 | 40 | | n | n | 38 | | 38 | 38 | n | 39/n | | n | 33 |
| | VM08 | 100 copies (Delta) | 33/35 | | 36 | 38 | 34 | | 38 | 34 | | 36 | | 35 | n | n | 36/37 | | 36 | 39 | | 38 | 36 | 38 | | 37 | 35 | n | 38/39 | | 37 | 35 |
| | VM13 | 100 copies (Delta) | 34/35 | | 35/36/36 | 37 | 36 | | 39 | 34 | | 35 | | 36 | 39 | 36 | 37/38 | | 38 | 38 | | 36 | 36 | 39 | | n | 36 | 35 | 39/38 | | 37 | 33 |
| | VM11 | 1,000 copies (Delta) | 31/31 | | 33 | 34 | 32 | | 37 | 30 | | 31 | | 32 | 36 | 32 | 34/34 | | 33 | 34 | | 33 | 32 | 33 | | 35 | 32 | 34 | 34/34 | | 33 | 29 |
| | VM07 | 1,000 copies (Omicron) | 31/32 | | 33 | 34 | 31 | | 35 | 29 | | 30 | | 31 | 34 | 31 | 33/33 | | 32 | 34 | | 31 | 32 | 33 | | 34 | 32 | 32 | 34/34 | | 33 | 29 |
| | VM14 | 1,000 copies (Omicron) | 31/31 | | 33 | 34 | 31 | | 35 | 29 | | 31 | | 31 | 35 | 31 | 33/34 | | 32 | 34 | | 32 | 32 | 33 | | 34 | 31 | 33 | 34/34 | | 33 | 29 |
| N2 | VM02 | blank | n/n | | n | n | n | | n | n | | n | | n | n | n | n/n | | n | n | | n | n | n | | n | n | n | n/n | | n | n |
| | VM04 | blank | n/n | | n | n | n | | n | n | | n | | n | n | n | n/n | | n | n | | n | n | n | | n | n | n | n/n | | n | n |
| | VM05 | Blank + FIP | n/n | | n | n | n | | n | n | | n | | n | n | n | n/n | | n | n | | n | n | n | | n | n | n | n/n | | n | n |
| | VM10 | Blank + FIP | n/n | | n | n | n | | n | n | | n | | n | n | n | n/n | | n | n | | n | n | n | | n | n | n | n/n | | n | n |
| | VM01 | 25 copies (Delta) | 35/n | | 37 | n | n | | 38 | 35 | | n | | 36 | n | n | n/n | | n | n | | 38 | 37 | n | | n | 36 | n | 39/39 | | n | n |
| | VM12 | 25 copies (Delta) | n/36 | | n | 40 | n | | n | n | | n | | 36 | n | 39 | 37/n | | 40 | n | | 38 | 37 | n | | 37 | 39 | n | n/n | | n | 39 |
| | VM03 | 50 copies (Delta) | 34/35 | | 37/n/n | 39 | 40 | | 40 | 37 | | 36 | | 37 | 42 | 39 | 39/n | | n | 41 | | 38 | 36 | n | | n | 38 | n | 39/n | | n | 34 |
| | VM06 | 50 copies (Delta) | 36/35 | | n | 38 | 37 | | n | 34 | | 35 | | 36 | n | n | 38/n | | 42 | --- | | 37 | 35 | 38 | | 38 | n | 38 | 41/n | | n | 36 |
| | VM09 | 50 copies (Delta) | 34/33 | | 37 | n | 39 | | n | 37 | | n | | 36 | n | 38 | n/37 | | 39 | 39 | | n | 37 | 40 | | 37 | 38 | n | 39/39 | | n | 35 |
| | VM08 | 100 copies (Delta) | 32/34 | | 36 | 40 | 39 | | 40 | 35 | | 36 | | 35 | n | 39 | 37/37 | | 40 | 38 | | 37 | n | 40 | | 38 | 37 | 38 | 40/41 | | 38 | 35 |
| | VM13 | 100 copies (Delta) | n/34 | | n/37/n | 37 | 36 | | 39 | 35 | | n | | 37 | 42 | 38 | 39/37 | | 39 | 41 | | 38 | 37 | 37 | | 38 | 37 | 39 | 39/40 | | 37 | 36 |
| | VM11 | 1,000 copies (Delta) | 30/31 | | 35 | 34 | 34 | | 38 | 32 | | 32 | | 32 | 42 | 33 | 35/34 | | 35 | 36 | | 33 | 33 | 35 | | 34 | 33 | 34 | 35/35 | | 35 | 31 |
| | VM07 | 1,000 copies (Omicron) | 30/30 | | 33 | 34 | 32 | | 36 | 28 | | 31 | | 31 | 36 | 32 | 33/33 | | 34 | 34 | | 32 | 32 | 34 | | 33 | 33 | 33 | 34/34 | | 34 | 31 |
| | VM14 | 1,000 copies (Omicron) | 30/30 | | 33 | 33 | 32 | | 35 | 30 | | 32 | | 31 | 40 | 32 | 32/33 | | 34 | 34 | | 31 | 32 | 33 | | 33 | 32 | 33 | 34/34 | | 33 | 31 |

Yellow color – false-negative results (Ct was not reported or value is above participant-specific cut off); pink color - false-positive results (Ct is below lab-specific cut-off); blue color – inconclusive result;
grey color – no result was submitted for the marker; n – no Ct value was reported; --- – no Ct value submitted for the particular sample; * - copy number in 50 µl used for RNA extraction. *Results from lab 337-3 is included for comparison purposes only and is excluded from statistical analysis.*

**Table S8: Ct values submitted by the laboratories for the N and N3 markers**

| Marker | Sample | Sample description | Participant Identification Number (PIN) | | | | | | | | | | | | | | | | | | | | | | | | | | | | | |
|---|---|---|---|---|---|---|---|---|---|---|---|---|---|---|---|---|---|---|---|---|---|---|---|---|---|---|---|---|---|---|---|---|
| | | | 8-6 | 11-3 | 32-10 | 86-4 | 238-2 | 287-2 | 312-2 | 334-8 | 335-11 | 337-3 | 338-17 | 340-7 | 341-2 | 342-5 | 343-4 | 346-8 | 347-2 | 349-10 | 352-4/5 | 357-8 | 359-9 | 366-9/10 | 367-8 | 368-1 | 373-4 | 891-5 | 892-2/3 | 893-2 | 894-2 | 895-1 |
| N | VM02 | blank | | n/n | | | | n | | | n | | | | | | | n | | | n | | | | | | | | | n/n | | |
| | VM04 | blank | | n/n | | | | n | | | n | | | | | | | n | | | n | | | | | | | | | n/n | | |
| | VM05 | Blank + FIP | | n/n | | | | n | | | n | | | | | | | n | | | n | | | | | | | | | n/n | | |
| | VM10 | Blank + FIP | | n | | | | n | | | n | | | | | | | n | | | n | | | | | | | | | n/n | | |
| | VM01 | 25 copies (Delta)* | | n/39 | | | | n | | | 35 | | | | | | | n | | | 36 | | | | | | | | | 35/35 | | |
| | VM12 | 25 copies (Delta) | | n/39 | | | | 37 | | | 40 | | | | | | | n | | | n | | | | | | | | | 36/36 | | |
| | VM03 | 50 copies (Delta) | | 33/34 | | | | 38 | | | 36 | | | | | | | n | | | 35 | | | | | | | | | 38/36 | | |
| | VM06 | 50 copies (Delta) | | n/37 | | | | 38 | | | 35 | | | | | | | n | | | 35 | | | | | | | | | 36/36 | | |
| | VM09 | 50 copies (Delta) | | 35/36 | | | | 35 | | | 38 | | | | | | | n | | | 36 | | | | | | | | | 35/36 | | |
| | VM08 | 100 copies (Delta) | | 35/27 | | | | 38 | | | 35 | | | | | | | 38 | | | 36 | | | | | | | | | 35/35 | | |
| | VM13 | 100 copies (Delta) | | 35/n | | | | 37 | | | 35 | | | | | | | 37 | | | 36 | | | | | | | | | 33/34 | | |
| | VM11 | 1,000 copies (Delta) | | 32/33 | | | | 33 | | | 31 | | | | | | | 34 | | | 30 | | | | | | | | | 31/31 | | |
| | VM07 | 1,000 copies (Omicron) | | 31/31 | | | | 32 | | | 31 | | | | | | | 33 | | | 30 | | | | | | | | | 31/31 | | |
| | VM14 | 1,000 copies (Omicron) | | 31/32 | | | | 32 | | | 31 | | | | | | | 32 | | | 31 | | | | | | | | | 30/31 | | |
| N3 | VM02 | blank | | | | | | | | | | | | | | | | | | | | | n | | | | | | | | | |
| | VM04 | blank | | | | | | | | | | | | | | | | | | | | | n | | | | | | | | | |
| | VM05 | Blank + FIP | | | | | | | | | | | | | | | | | | | | | n | | | | | | | | | |
| | VM10 | Blank + FIP | | | | | | | | | | | | | | | | | | | | | n | | | | | | | | | |
| | VM01 | 25 copies (Delta) | | | | | | | | | | | | | | | | | | | | | 36 | | | | | | | | | |
| | VM12 | 25 copies (Delta) | | | | | | | | | | | | | | | | | | | | | n | | | | | | | | | |
| | VM03 | 50 copies (Delta) | | | | | | | | | | | | | | | | | | | | | 36 | | | | | | | | | |
| | VM06 | 50 copies (Delta) | | | | | | | | | | | | | | | | | | | | | 37 | | | | | | | | | |
| | VM09 | 50 copies (Delta) | | | | | | | | | | | | | | | | | | | | | 35 | | | | | | | | | |
| | VM08 | 100 copies (Delta) | | | | | | | | | | | | | | | | | | | | | 35 | | | | | | | | | |
| | VM13 | 100 copies (Delta) | | | | | | | | | | | | | | | | | | | | | n | | | | | | | | | |
| | VM11 | 1,000 copies (Delta) | | | | | | | | | | | | | | | | | | | | | 34 | | | | | | | | | |

| | | | | | | | | | | | | | | | | | | | | | | | |
|---|---|---|---|---|---|---|---|---|---|---|---|---|---|---|---|---|---|---|---|---|---|---|---|
| | VM07 | 1,000 copies (Omicron) | | | | | | | | | | | | | | | | | | | | | 32 |
| | VM14 | 1,000 copies (Omicron) | | | | | | | | | | | | | | | | | | | | | 32 |

Yellow color – false-negative results (Ct was not reported or value above participant-specific cut off); blue color – inconclusive result; grey color – no result was submitted for the marker; n – no Ct value was reported; * - copy number in 50 μl used for RNA extraction.

**Table S9: Ct values submitted by the laboratories for the E and RdRP markers**

| Marker | Sample | Sample Description | Participant Identification Number (PIN) | | | | | | | | | | | | | | | | | | | | | | | | | | | | | |
|---|---|---|---|---|---|---|---|---|---|---|---|---|---|---|---|---|---|---|---|---|---|---|---|---|---|---|---|---|---|---|---|---|
| | | | 8-6 | 11-3 | 32-10 | 86-4 | 238-2 | 287-2 | 312-2 | 334-8 | 335-11 | 337-3 | 338-17 | 340-7 | 341-2 | 342-5 | 343-4 | 346-8 | 347-2 | 349-10 | 352-4/5 | 357-8 | 359-9 | 366-9/10 | 367-8 | 368-1 | 373-4 | 891-5 | 892-2/3 | 893-2 | 894-2 | 895-1 |
| E | VM02 | blank | | | | | | | | | | | n | | | | | | | | | | | | | | | | | | | |
| | VM04 | blank | | | | | | | | | | | n | | | | | | | | | | | | | | | | | | | |
| | VM05 | Blank + FIP | | | | | | | | | | | n | | | | | | | | | | | | | | | | | | | |
| | VM10 | Blank + FIP | | | | | | | | | | | n | | | | | | | | | | | | | | | | | | | |
| | VM01 | 25 copies (Delta)* | | | | | | | | | | | 35 | | | | | | | | | | | | | | | | | | | |
| | VM12 | 25 copies (Delta) | | | | | | | | | | | 33 | | | | | | | | | | | | | | | | | | | |
| | VM03 | 50 copies (Delta) | | | | | | | | | | | 35 | | | | | | | | | | | | | | | | | | | |
| | VM06 | 50 copies (Delta) | | | | | | | | | | | 34 | | | | | | | | | | | | | | | | | | | |
| | VM09 | 50 copies (Delta) | | | | | | | | | | | 32 | | | | | | | | | | | | | | | | | | | |
| | VM08 | 100 copies (Delta) | | | | | | | | | | | 33 | | | | | | | | | | | | | | | | | | | |
| | VM13 | 100 copies (Delta) | | | | | | | | | | | 34 | | | | | | | | | | | | | | | | | | | |
| | VM11 | 1,000 copies (Delta) | | | | | | | | | | | 28 | | | | | | | | | | | | | | | | | | | |
| | VM07 | 1,000 copies (Omicron) | | | | | | | | | | | 29 | | | | | | | | | | | | | | | | | | | |
| | VM14 | 1,000 copies (Omicron) | | | | | | | | | | | 29 | | | | | | | | | | | | | | | | | | | |
| RdRP | VM02 | blank | | | | | | | | | | | n | | | | | | | | | | | | 42 | | | | | | | |
| | VM04 | blank | | | | | | | | | | | n | | | | | | | | | | | | n | | | | | | | |
| | VM05 | Blank + FIP | | | | | | | | | | | n | | | | | | | | | | | | n | | | | | | | |
| | VM10 | Blank + FIP | | | | | | | | | | | n | | | | | | | | | | | | n | | | | | | | |
| | VM01 | 25 copies (Delta) | | | | | | | | | | | 38 | | | | | | | | | | | | n | | | | | | | |
| | VM12 | 25 copies (Delta) | | | | | | | | | | | 34 | | | | | | | | | | | | n | | | | | | | |
| | VM03 | 50 copies (Delta) | | | | | | | | | | | n | | | | | | | | | | | | n | | | | | | | |
| | VM06 | 50 copies (Delta) | | | | | | | | | | | 34 | | | | | | | | | | | | 42 | | | | | | | |
| | VM09 | 50 copies (Delta) | | | | | | | | | | | 35 | | | | | | | | | | | | 43 | | | | | | | |
| | VM08 | 100 copies (Delta) | | | | | | | | | | | 35 | | | | | | | | | | | | 41 | | | | | | | |
| | VM13 | 100 copies (Delta) | | | | | | | | | | | 34 | | | | | | | | | | | | n | | | | | | | |
| | VM11 | 1,000 copies (Delta) | | | | | | | | | | | 29 | | | | | | | | | | | | n | | | | | | | |

| | | | | | | | | | | | | | | | | | | | | | | | | | | |
|---|---|---|---|---|---|---|---|---|---|---|---|---|---|---|---|---|---|---|---|---|---|---|---|---|---|---|
| | VM07 | 1,000 copies (Omicron) | | | | | | | | | | | 31 | | | | | | | | | | | | | n |
| | VM14 | 1,000 copies (Omicron) | | | | | | | | | | | 31 | | | | | | | | | | | | | 41 |

Yellow color – false-negative results (Ct was not reported or value was above participant-specific cut off); pink color - false-positive result (Ct was below the lab-specific cut-off value);
grey color – no result was submitted for the marker; n – no Ct value was reported; * - copy number in 50 µL used for RNA extraction.

**Table S10: Ct values submitted by the laboratories for the ORF1ab and S markers**

| Marker | Sample | Sample Description | Participant Identification Number (PIN) | | | | | | | | | | | | | | | | | | | | | | | | | | | | | |
|---|---|---|---|---|---|---|---|---|---|---|---|---|---|---|---|---|---|---|---|---|---|---|---|---|---|---|---|---|---|---|---|---|
| | | | 8-6 | 11-3 | 32-10 | 86-4 | 238-2 | 287-2 | 312-2 | 334-8 | 335-11 | 337-3 | 338-17 | 340-7 | 341-2 | 342-5 | 343-4 | 346-8 | 347-2 | 349-10 | 352-4/5 | 357-8 | 359-9 | 366-9/10 | 367-8 | 368-1 | 373-4 | 891-5 | 892-2/3 | 893-2 | 894-2 | 895-1 |
| | VM02 | blank | | n/n | | | | | | | n | | | | | | | n | | | n | | | | | | | | | | | |
| | VM04 | blank | | n/n | | | | | | | n | | | | | | | n | | | n | | | | | | | | | | | |
| | VM05 | Blank + FIP | | n/n | | | | | | | n | | | | | | | n | | | n | | | | | | | | | | | |
| | VM10 | Blank + FIP | | n | | | | | | | n | | | | | | | n | | | n | | | | | | | | | | | |
| | VM01 | 25 copies (Delta)* | | n/n | | | | | | | 35 | | | | | | | n | | | 33 | | | | | | | | | | | |
| | VM12 | 25 copies (Delta) | | 37/n | | | | | | | 35 | | | | | | | n | | | n | | | | | | | | | | | |
| | VM03 | 50 copies (Delta) | | n/37 | | | | | | | 34 | | | | | | | n | | | 34 | | | | | | | | | | | |
| ORF1ab | VM06 | 50 copies (Delta) | | n/n | | | | | | | 36 | | | | | | | 38 | | | n | | | | | | | | | | | |
| | VM09 | 50 copies (Delta) | | 38/36 | | | | | | | 35 | | | | | | | 38 | | | 34 | | | | | | | | | | | |
| | VM08 | 100 copies (Delta) | | 36/35 | | | | | | | 35 | | | | | | | 37 | | | 33 | | | | | | | | | | | |
| | VM13 | 100 copies (Delta) | | 36/36 | | | | | | | 34 | | | | | | | 37 | | | 33 | | | | | | | | | | | |
| | VM11 | 1,000 copies (Delta) | | 33/33 | | | | | | | 31 | | | | | | | 34 | | | 30 | | | | | | | | | | | |
| | VM07 | 1,000 copies (Omicron) | | 32/32 | | | | | | | 32 | | | | | | | 33 | | | 30 | | | | | | | | | | | |
| | VM14 | 1,000 copies (Omicron) | | 32/32 | | | | | | | 32 | | | | | | | 34 | | | 30 | | | | | | | | | | | |
| | VM02 | blank | | n/n | | | | | | | n | | | | | | | n | | | n | | | | | | | | | | | |
| | VM04 | blank | | n/n | | | | | | | n | | | | | | | n | | | n | | | | | | | | | | | |
| | VM05 | Blank + FIP | | n/n | | | | | | | n | | | | | | | n | | | n | | | | | | | | | | | |
| | VM10 | Blank + FIP | | n | | | | | | | n | | | | | | | n | | | n | | | | | | | | | | | |
| S | VM01 | 25 copies (Delta) | | 38/n | | | | | | | 36 | | | | | | | n | | | 37 | | | | | | | | | | | |
| | VM12 | 25 copies (Delta) | | n/n | | | | | | | 32 | | | | | | | n | | | n | | | | | | | | | | | |
| | VM03 | 50 copies (Delta) | | 37/36 | | | | | | | 37 | | | | | | | n | | | 35 | | | | | | | | | | | |
| | VM06 | 50 copies (Delta) | | 38/n | | | | | | | 36 | | | | | | | n | | | n | | | | | | | | | | | |

| | | | | | | | | | | | | | | | | | | | | | |
|---|---|---|---|---|---|---|---|---|---|---|---|---|---|---|---|---|---|---|---|---|---|
| | VM09 | 50 copies (Delta) | | n/n | | | | | | | 36 | | | | | | | 39 | | | 34 |
| | VM08 | 100 copies (Delta) | | 35/38 | | | | | | | 34 | | | | | | | n | | | 34 |
| | VM13 | 100 copies (Delta) | | n/n | | | | | | | 34 | | | | | | | 37 | | | 35 |
| | VM11 | 1,000 copies (Delta) | | 33/33 | | | | | | | 31 | | | | | | | 34 | | | 31 |
| | VM07 | 1,000 copies (Omicron) | | n/n | | | | | | | n | | | | | | | n | | | n |
| | VM14 | 1,000 copies (Omicron) | | n/n | | | | | | | n | | | | | | | n | | | n |

Yellow color – false-negative results (Ct was not reported or value above participant-specific cut off); blue color – inconclusive result; grey color – no result was submitted for the marker; n – no Ct value was reported; * - copy number in 50 µl used for RNA extraction.
